# Premorbid frailty predicts short and long term outcomes of reperfusion treatment in acute stroke

**DOI:** 10.1101/2021.11.28.21266639

**Authors:** Andrea Pilotto, Cora Brass, Klaus Fassbender, Fatma Merzou, Andrea Morotti, Niklas Kämpfer, A Antonio Siniscalchi, A Alessandro Padovani, Piergiorgio Lochner

**Author notes:** **Corresponding author:** Andrea Pilotto, MD, Neurology Unit, Department of Clinical and Experimental Sciences, University of Brescia, P.zale Spedali Civili, 1 - 25123 Brescia, Italy.

## Abstract

**Background:** Frailty is the most important short and long term predictor of disability in the elderly and thus might influence the clinical outcome of acute treatment of stroke.

**Objective:** to evaluate whether frailty predicts short- and long term all-cause mortality and neurological recovery in elderly patients who underwent reperfusion acute treatment of stroke.

**Methods:** the study included consecutive patients older than 65 years who underwent reperfusion treatment in a single stroke Unit from 2015 to 2016. Predictors of stroke outcomes were assessed including demographics, baseline NIHSS, time to needle, treatment and medical complications. Premorbid Frailty was assessed with a comprehensive geriatric assessment (CGA) including functional, nutritional, cognitive, social and comorbidities status. At three and twelve months, all-cause death and clinical recovery (using modified Ranking scale, mRS) were evaluated.

**Results:** One-hundred and two patients who underwent acute reperfusion treatment for stroke entered the study (mean age 77.5, 65-94 years). Frailty was diagnosed in 32 out of 70 patients and associated with older age (p=0.001) but no differences in baseline NIHSS score, vascular risk profile or treatment management strategy. Frailty status was associated with worse improvement at 24 hours and higher in-hospital mortality. At follow-up, frail patients showed poorer survival at 3 (25% vs 3%, p=0.008) and 12 (38% vs 7%, p=0.001) months. Frailty was the best predictor of neurological recovery at one year follow-up (mRS 3.2 ± 1.9 vs 1.9 ± 1.9).

**Discussion:** frailty is an important predictor of efficacy of acute treatment of stroke beyond classical predictors of stroke outcomes. Larger longitudinal studies are thus warranted in order to evaluate the risk-benefit of reperfusion treatment in the growing elderly frail population.

## INTRODUCTION

Stroke is the fourth cause of death and the leading cause of disability worldwide[1,2]. The main driver for increased stroke incidence and prevalence is ageing of the population. Older age is at the same time an important risk factor for stroke and the most relevant predictor of worse outcome and increased disability after an ischemic cerebral event[3,4]. Given that the number of incident stroke in the geriatric population is expected to rapidly increase, understanding what underlying factors portend the response to the medical management of stroke and reperfusion treatments is of critical interest. In fact, best-evidence based strategies for stroke treatment and prevention are largely not validated in the older population and considerable gaps of knowledge exist in this specific population[1,5].

Frailty is a common geriatric condition characterized by decreased resilience to stressor and an association with increased risk of mortality and disability[6]. Previous studies found that frailty might impact on survival and recovery after ischemic stroke[4] or transient ischemic attack[7] in the elderly population, independently from age and the cumulative number of vascular risk factors.

To date, however, no studies specifically evaluated the impact of premorbid frailty in the response of acute reperfusion treatments of stroke in elderly patients. This unexplored issue is pivotal in order to plan the medical resources and for the developing of specific guidelines for stroke management in the elderly population[1]. In this study, we specifically hypothesize that premorbid frailty is an important predictor of short and long term poor response to reperfusion treatment in the elderly, independently from age and brain-related factors.

## MATERIALS AND METHODS

### Recruitment and inclusion criteria

All patients consecutively admitted to the Stroke Unit Department of Neurology, University of the Saarland, Homburg, Saar, Germany between Jan 1 2015 and December 31, 2018, with symptoms suggestive of acute stroke were screened. Inclusion criteria were: (1) age above 65 years; (2) diagnosis of acute stroke and CT/MRI scan results with no evidence of other causes that might explain the neurologic deficits; (3) reperfusion treatment with rtPA (alone or combined with thrombectomy); (4) informed consent approval for the retrospective use of clinical data and follow-up at one year; (5) a complete availability of data for the calculation of premorbid frailty measure at baseline.

All patients received the treatment based on established guidelines^1^ and all cerebral large-artery occlusion was confirmed by head and neck CT angiography. Patients received intravenous thrombolysis alone or combined treatment with ‘‘contemporary/as soon as possible’’ endovascular mechanical thrombectomy^1^. The Institutional Ethical Standards Committee on human experimentation at Saarland Hospital provided approval for the study 269/17).

### Data Collection

Epidemiological, demographical, clinical, treatment, and outcome data were extracted from both printed and electronic medical records using standardized anonymized data collection forms. All data were imputed and checked by three physicians (KF, FR, PGL). We systematically assessed the following variables: Stroke severity by the National Institutes of Health Stroke Scale score^8^ at onset, 24 hours after onset, time from stroke symptoms onset to treatment, total days of hospitalisation, ranking scale at discharge.

#### Frailty assessment

In order to calculate the MPI a comprehensive geriatric assessment (CGA) including eight domains (comorbidity index, number of drugs, pressure sores, dependency on basic and instrumental activities of daily living, cognitive, nutritional and social status) was applied[9]. The sum of scores of each domain was divided by eight to obtain a final MPI risk score between 0 = no risk and 1 = higher risk of mortality (Supplementary for detailed cut off values for each domain). The patients were thus dichotomized as robust (MPI value <0.33) and frail (MPI value ≥ 0.34) and the MPI was used as continuous variable for linear regression analyses.

#### Outcome measures

In hospital complications included pneumonia, deep venous thrombosis and in-hospital death. A telemedicine follow-up at three months and twelve months was additionally applied to all patients in order to evaluate the survival and long-term mRS scale.

### Statistical analysis

Continuous and categorical variables are reported as mean and standard deviation and percentage (n) respectively. Differences between patients with different acute treatments (rtPA alone vs thrombectomy plus rtPA) and between robust and frail patients were compared by Mann-Whitney U test, χ^2^ test or Fisher’s exact test where appropriate. To explore the risk factors associated with modified Ranking scale univariable and multivariable logistic regression models were implemented. For multivariate analysis, to avoid overfitting in the model, variables were chosen based on previous findings and included age, sex, baseline NIHSS, time to needle, treatment, vascular risk factors as predictors. For death, a cox-regression including the same variables was implemented. Data analyses were carried out using SPSS software (version 22.0).

## RESULTS

Two-hundred ten patients older than 65 years underwent reperfusion treatments between January 1^st^, 2015 and December 31rd 2018 were consecutively screened. Of them, 102 subjects agreed to the participation of the study including the complete frailty premorbid assessment and the clinical follow-up (mean age 77.3 ± 6.7, range 65-94). Selected patients were comparable for demographics and clinical variables with patients who refused the participation (Supplementary Figure 1 and supplementary Table 1). Seventy-one patients were treated with fibrinolysis alone, while 31 patients additionally underwent a thrombectomy for large vessel occlusion. The different treatment groups were comparable for age, sex, vascular risk factors distribution exception made for atrial fibrillation, which was more common in patients who underwent thrombectomy (Table 1). The treatment groups, as expected, differ for time to needle, baseline and 24-hours NIHSS score.

**Table 1.** Clinical and demographic characteristics of patients included according to the treatment. ^$^ Comparison between Robust and frail subgroups have been performed by Mann-Whitney *U* test or Fisher exact test for continuous and dichotomous variables, respectively.

|  | All | Thrombectomy plus rtPA | rtPA only | P |
| --- | --- | --- | --- | --- |
| n | 102 | 31 | 71 |  |
| Age, years | 77.2 ± 6.7 | 76.6 ± 6.8 | 77.5 ± 6.8 | 0.45 |
| Sex, f % | 42.2 (43) | 48.4 (15) | 40.0 (28) | 0.28 |
| NIHSS admission | 9.2 ± 6.6 | 12.4 | 7.70 ± 5.5 | **0.001** |
| NIHSS 24 hours | 7.9 ± 10.8 | 13.4 | 5.5 ± 6.8 | **0.018** |
| Time to needle, min | 94 ± 38 | 84 ± 40 | 99 ± 36 | **0.009** |
| **Vascular risk factors** |  |  |  |  |
| Hypertension | 83.2 (84) | 80.6 (25) | 84.3 (59) | 0.42 |
| Diabetes | 35.2 (36) | 29 (9) | 38.6 (43) | 0.24 |
| Dyslipidemia | 31.4 (32) | 25.8 (8) | 32.4 (23) | 0.32 |
| Smoking history | 15.8 (16) | 19.4 (6) | 14.3 (10) | 0.35 |
| Ischemic heart disease | 17.6 (18) | 19.4 (6) | 15.4 (11) | 0.42 |
| Atrial fibrillation | 15.8 (16) | 45.2 (14) | 19.7 (14) | **0.002** |
| **CGA assessment** |  |  |  |  |
| ADL | 5.4 ± 1.4 | 5.7 ± 0.9 | 5.2 1.6 | 0.13 |
| IADL | 6.1 ± 2.6 | 6.0 ± 2.6 | 6.3 2.4 | 0.74 |
| MNA | 12.6 ± 2.1 | 12.5 ± 2.0 | 12.6 2.1 | 0.73 |
| ESS, score | 18.5 ± 2.3 | 18.6 ± 2.1 | 18.4 2.4 | 0.72 |
| CIRS-CI | 1.8 ± 1.3 | 1.6 ± 1.4 | 1.8 1.3 | 0.26 |
| Number of medications | 5.5 ± 3.9 | 6.1 ± 3.8 | 5.3 3.9 | 0.27 |
| Cognition, dementia % | 11.9 (12) | 10 (12.8) | 6.8 (2) | 0.07 |
| Social conditions, alone % | 17.8 (18) | 16.1 (5) | 18.6 (13) | 0.24 |
| MPI | 0.24 0.18 | 0.22 ± 0.16 | 0.25 0.19 | 0.76 |
| Frail , dichotomic | 30 (31) | 31 (22) | 29 (9) | 1.00 |

**Abbreviations:** ADL, activities of daily living; CIRS-CI, Cumulative Illness Rating scale-comorbidity index; ESS, Exton smith sores scale, f, female; IADL, instrumental activities of daily living; min, minutes; MNA, mininutritional assessment, MPI, multidimensional prognostic index; NIHSS, National Institutes of Health Stroke Scale y, years

### Frailty assessment, stroke presentation and short term outcomes

The CGA assessment identified 32 frail and 70 robust patients (Table 2). Frail patients were older compared to robust subgroup (80.5 ± 6.2 vs 75.8 ± 6.4 years) with similar distribution of sex, vascular risk factors and reperfusion management strategies. According to CGA classification, the scores of each domain of frailty differ between robust and frail subgroups (p<0.005). Baseline NIHSS was similar between frail and robust patients while 24-hour NIHSS-after reperfusion treatment differ between groups (p=0.016). Frail patients exhibited higher in hospital mortality (9.4% vs 0%, p=0.029) and deep venous thrombosis but similar rate of pneumonia and total days of hospitalization compared to robust patients. After exclusion of deceased patients, linear regression analyses identified age (T 2.7, p=0.008,) and Diabetes (T 2.4, p=0.015) as predictor of mRS adjusting for sex, baseline NIHSS, vascular risk factors, frail status, treatment adopted and time door to needle.

**Table 2.**
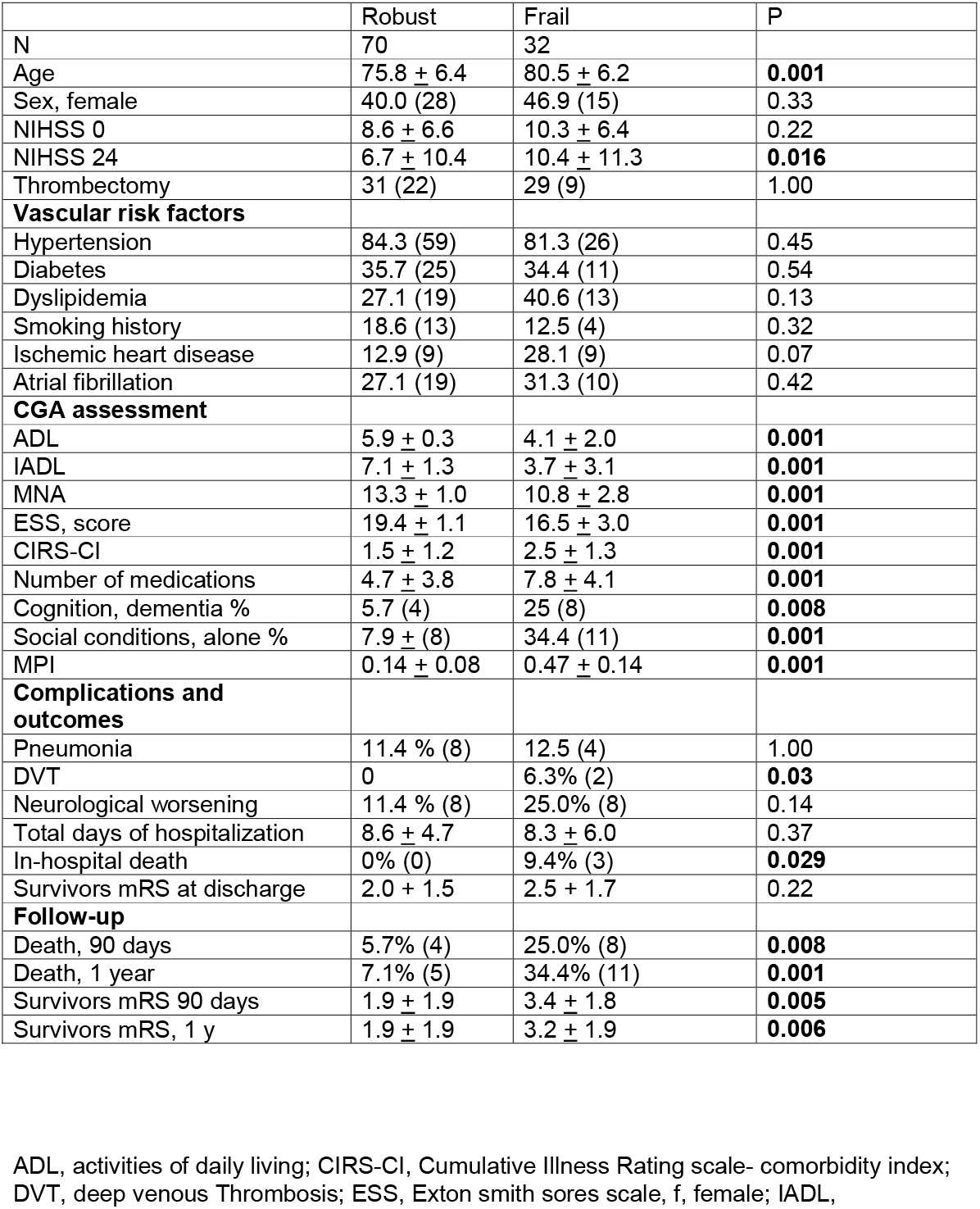
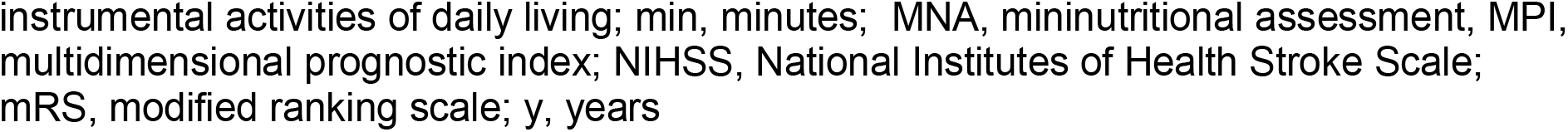
Clinical and demographic characteristics of patients according to frail and robust classification. ^$^ Comparison between Robust and frail subgroups have been performed by Mann-Whitney *U* test or Fisher exact test for continuous and dichotomous variables, respectively.

|  | Robust | Frail | P |
| --- | --- | --- | --- |
| N | 70 | 32 |  |
| Age | 75.8 ± 6.4 | 80.5 ± 6.2 | **0.001** |
| Sex, female | 40.0 (28) | 46.9 (15) | 0.33 |
| NIHSS 0 | 8.6 ± 6.6 | 10.3 ± 6.4 | 0.22 |
| NIHSS 24 | 6.7 ± 10.4 | 10.4 ± 11.3 | **0.016** |
| Thrombectomy | 31 (22) | 29 (9) | 1.00 |
| **Vascular risk factors** |  |  |  |
| Hypertension | 84.3 (59) | 81.3 (26) | 0.45 |
| Diabetes | 35.7 (25) | 34.4 (11) | 0.54 |
| Dyslipidemia | 27.1 (19) | 40.6 (13) | 0.13 |
| Smoking history | 18.6 (13) | 12.5 (4) | 0.32 |
| Ischemic heart disease | 12.9 (9) | 28.1 (9) | 0.07 |
| Atrial fibrillation | 27.1 (19) | 31.3 (10) | 0.42 |
| **CGA assessment** |  |  |  |
| ADL | 5.9 ± 0.3 | 4.1 ± 2.0 | **0.001** |
| IADL | 7.1 ± 1.3 | 3.7 ± 3.1 | **0.001** |
| MNA | 13.3 ± 1.0 | 10.8 ± 2.8 | **0.001** |
| ESS, score | 19.4 ± 1.1 | 16.5 ± 3.0 | **0.001** |
| CIRS-CI | 1.5 ± 1.2 | 2.5 ± 1.3 | **0.001** |
| Number of medications | 4.7 ± 3.8 | 7.8 ± 4.1 | **0.001** |
| Cognition, dementia % | 5.7 (4) | 25 (8) | **0.008** |
| Social conditions, alone % | 7.9 ± (8) | 34.4 (11) | **0.001** |
| MPI | 0.14 ± 0.08 | 0.47 ± 0.14 | **0.001** |
| **Complications and outcomes** |  |  |  |
| Pneumonia | 11.4 % (8) | 12.5 (4) | 1.00 |
| DVT | 0 | 6.3% (2) | **0.03** |
| Neurological worsening | 11.4 % (8) | 25.0% (8) | 0.14 |
| Total days of hospitalization | 8.6 ± 4.7 | 8.3 ± 6.0 | 0.37 |
| In-hospital death | 0% (0) | 9.4% (3) | **0.029** |
| Survivors mRS at discharge | 2.0 ± 1.5 | 2.5 ± 1.7 | 0.22 |
| **Follow-up** |  |  |  |
| Death, 90 days | 5.7% (4) | 25.0% (8) | **0.008** |
| Death, 1 year | 7.1% (5) | 34.4% (11) | **0.001** |
| Survivors mRS 90 days | 1.9 ± 1.9 | 3.4 ± 1.8 | **0.005** |
| Survivors mRS, 1 y | 1.9 ± 1.9 | 3.2 ± 1.9 | **0.006** |

ADL, activities of daily living; CIRS-CI, Cumulative Illness Rating scale- comorbidity index; DVT, deep venous Thrombosis; ESS, Exton smith sores scale, f, female; IADL,
instrumental activities of daily living; min, minutes; MNA, mininutritional assessment, MPI, multidimensional prognostic index; NIHSS, National Institutes of Health Stroke Scale; mRS, modified ranking scale; y, years

### Frailty impact on death and disability at follow-up

In Cox-regression analyses the frail condition (MPI) emerged as best predictor of 12 months death in stroke patients (p=0.008, Wald 7.1), followed by diabetes (p=0.014, Wald 6.1) and age (p=0.031, Wald 4.6) after adjusting for the effect of sex, reperfusion treatment, baseline NIHSS, hypertension, atrial fibrillation and dyslipidemia (Figure 1). After exclusion of death cases, linear regression analyses identified MPI (p=0.001), age (p=0.001) and baseline NIHSS (p=0.013) as best predictors of mRS in patients treated with rtPA only at 12 months. In patients who underwent thrombectomy, MPI was the only predictor of mRS at 12 months (p=0.016) adjusting for the effect of age, sex, baseline NIHSS and vascular comorbidities.

**Figure 1.**
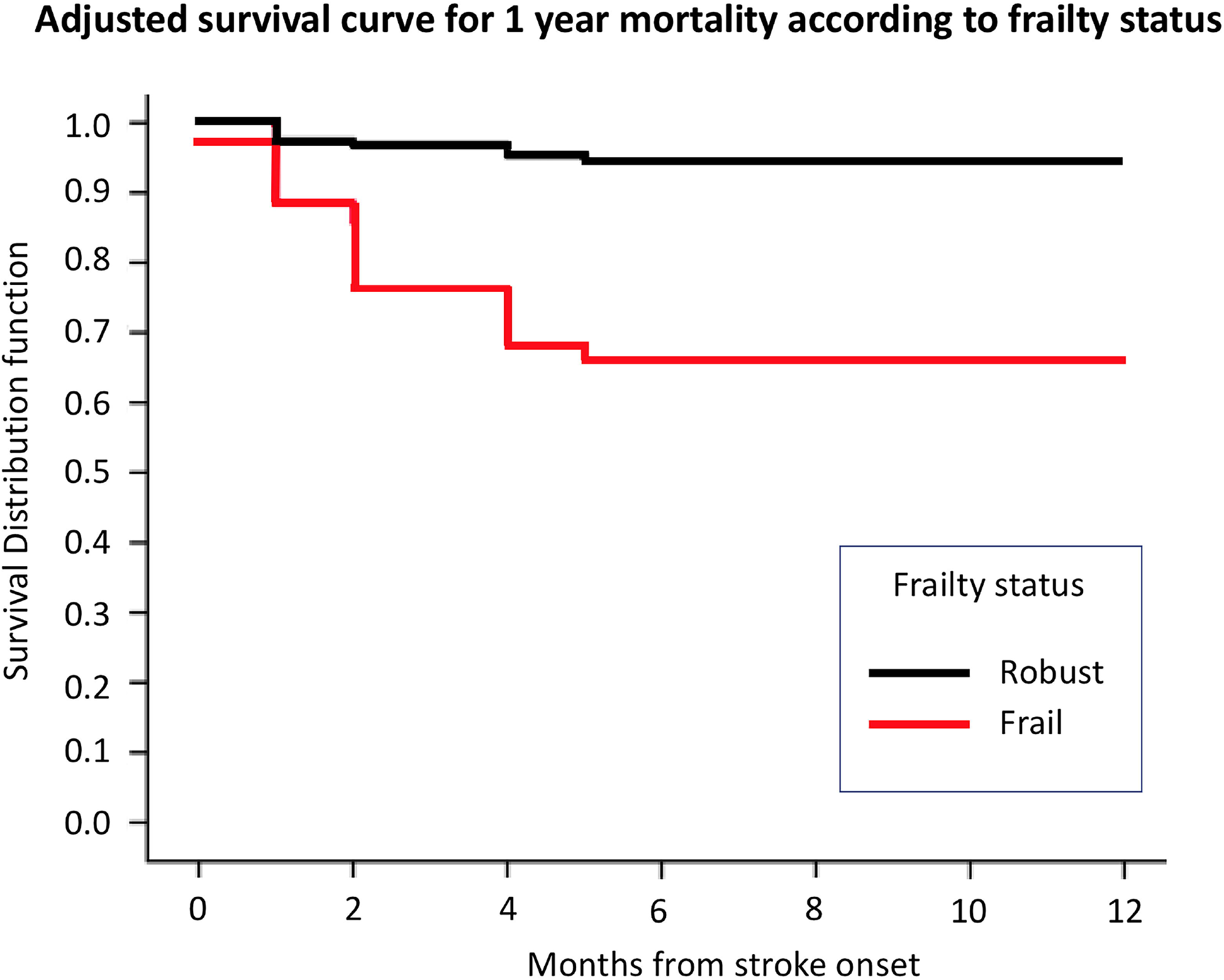
Survival at 12-months after the hospitalization for acute stroke according to Frail and Robust classification

## DISCUSSION

Our results demonstrate that premorbid frailty is a major determinant of the short and long-term response to acute reperfusion treatment in elderly patients with stroke. Notably, age is a major risk factor for stroke and the number of stroke in elderly patients is expected to rapidly increase in the next decade[4]. Despite its strong influence on case fatality, however, older age alone is still poor predictor of functional recovery in elderly patients[3,10,11]. Frailty is the most important predictor of disability and death in the ageing population and we hypothesize it can play a major role in explaining the wide variability of outcomes observed[4,12]. In this study, we evaluated premorbid frailty status and brain-specific predictors in consecutive patients who underwent a reperfusion treatment and hospitalized for stroke. As frailty measure we adopted the MPI score, one of best prognostic index for older adults[6] largely validated in different settings[7,9]. The index incorporate eight domains associated with worse outcomes in the elderly, including the number and severity of comorbidities, the functional, cognitive, nutritional and social status. In the study, frail patients were similar in terms of stroke severity, vascular risk factor profile, reperfusion treatment adopted but exhibited higher vulnerability in all geriatric domains assessed. In the short-term, frailty status was associated with lower improvement at the NIHSS scale, higher incidence of deep venous thrombosis and death. In the long-term, frailty emerged as the best predictor of survival and disability recovery at 3 and 12 months independently from age, reperfusion treatment adopted and vascular risk factor profile.

These findings are of great importance, as they might explain the large quote of futile recanalization treatment and wide variability of short and long-term outcomes observed in clinical practice[4,12,13].

Strengths of this study were the accurate selection criteria, the use of an extensive and validated measure of frailty and the long-term follow-up. However, the limited sample size did not allow a separate subanalyses evaluating the impact of frailty with a stratification of patients according to the severity and localization of occlusion[13,15]. Second, frailty might represent a selection bias conditioning the access to reperfusion treatment a priori. In our sample, interestingly, frail patients underwent a similar pre-hospital and in-hospital management but further studies including patients who did not undergo reperfusion are definitively needed to confirm these findings. Third, the small numbers also limited evaluation of single subdomains of frailty mostly associated with poor outcomes, an issue of major importance for implementing a shorter version of the MPI score for acute management setting.

Limitations notwithstanding, this is the first study showing that frailty play a major role in predicting the short and long term outcomes in elderly treated stroke patients. This is pivotal for an adequate management of stroke in the elderly and should definitively stimulate larger multicenter trials in order to evaluate the single-subject and healthcare systems implication of frailty stratification of stroke in older adults.

## Data Availability

All data produced in the present study are available upon reasonable request to the authors

## STATEMENTS AND DECLARATIONS

**Competing interest statement regarding the submitted work: none**

**Competing interest statement outside the submitted work**

Andrea Pilotto is supported by IMI H2020 initiative (MI2-2018-15-06) paid to the university of Brescia Italian Ministry of Health paid to the university of Brescia; he received Lectures honoraria from Bial, Biomarin, Abbvie, CHiesi and Zambon pharmaceuticals-paymemnt made to A.P. as an individual; he received research support from Bial, Biomarin, Abbvie, CHiesi and Zambon pharmaceuticals-payment made to the Institution University of Brescia,

Klaus Fassbender, Fatma Merzou, Andrea Morotti, Niklas Kämpfer and Antonio Siniscalchi have no conflict of interests

Alessandro Padovani received grant support from Ministry of Health (MINSAL) and Ministry of Education, Research and University (MIUR), from CARIPLO Foundation; personal compensation as a consultant/scientific advisory board member for Biogen 2019-2020-2021 Roche 2019-2020 Nutricia 2020-2021 General Healthcare (GE) 2019; he received honoraria for lectures at meeting ADPD2020 from Roche, Lecture at meeting of the Italian society of Neurology 2020 from Biogen and from Roche, Lecture at meeting AIP 2020 and 2021 from Biogen and from Nutricia, Educational Consulting 2019-2020-2021 from Biogen

Piergiorgio Lochner has no conflict of interest

