## supplementary material for "Premorbid frailty predicts short and long term outcomes of reperfusion treatment in acute stroke"

**Supplementary Table 1**

Clinical and demographic characteristics of patients included and excluded from the study

|  | included | excluded | P |
| --- | --- | --- | --- |
| n | 102 | 109 |  |
| Age, years | 77.2 ± 6.7 | 78.3 ± 6.9 | 0.29 |
| Sex, f % | 42.2 (43) | 45.0 (49) | 0.39 |
| NIHSS admission | 9.2 ± 6.6 | 11.8 ± 10.2 | **0.02** |
| **Vascular risk factors** |  |  |  |
| Hypertension | 83.3 (84) | 82.6 (90) | 0.52 |
| Diabetes | 35.3 (36) | 38.5 (42) | 0.36 |
| Dyslipidemia | 31.4 (32) | 33.9 (37) | 0.40 |
| Smoking history | 15.8 (16) | 14.3 (10) | 0.35 |
| Ischemic heart disease | 17.6 (18) | 21.1 (23) | 0.32 |
| Atrial fibrillation | 28.4 (29%) | 33.9 (37) | 0.23 |

**Abbreviations:** ADL, activities of daily living; CIRS-CI, Cumulative Illness Rating scale- comorbidity index; ESS, Exton smith sores scale, f, female; IADL, instrumental activities of daily living; min, minutes; MNA, mininutritional assessment, MPI, multidimensional prognostic index; y, years

**Supplementary Table 2**

| **a) Model for mRS at 12 months without MPI (R^2^ 0.42)** | | |  |  |  |
| --- | --- | --- | --- | --- | --- |
| **Independent variables** | **B** | **Standard error** | **Beta** | **t** | **p-value** |
| **Constant** | -7.383 | 2.32 |  | -3.196 | 0.002 |
| **age** | 0.102 | 0.029 | 0.296 | 3.442 | 0.001 |
| **Gender** | 1.191 | 0.994 | 0.195 | 1.198 | 0.239 |
| **Baseline NIHSS** | 0.089 | 0.025 | 0.307 | 3.605 | 0.001 |
| **Treatment** | 1.054 | 0.451 | 0.208 | 2.337 | 0.021 |
| **Time to needle** | 0.007 | 0.005 | 0.112 | 1.484 | 0.14 |
| **Diabetes** | 0.725 | 0.400 | 0.146 | 1.814 | 0.073 |
| **Hypertension** | -0.226 | 0.508 | -0.035 | -0.445 | 0.657 |
| **Atrial fibrillation** | 0.387 | 0.423 | 0.074 | 0.914 | 0.363 |
| **Dyslipidemia** | 0.151 | 0.411 | 0.029 | 0.367 | 0.715 |
| **b) Model for mRS at 12 months without MPI (R^2^ 0.51)** | | | | | |
| **Constant** | -7.383 | 2.32 |  | -3.196 | 0.002 |
| **age** | 0.102 | 0.029 | 0.296 | 3.442 | 0.001 |
| **Gender** | -0.254 | 0.375 | -0.055 | -0.678 | 0.500 |
| **Baseline NIHSS** | 0.074 | 0.029 | 0.212 | 2.564 | 0.012 |
| **Treatment** | 0.887 | 0.420 | 0.178 | 2.111 | 0.038 |
| **Time to needle** | 0.007 | 0.005 | 0.111 | 1.404 | 0.164 |
| **MPI score** | 4.374 | 1.094 | 0.350 | 3.999 | 0.001 |
| **Diabetes** | 0.462 | 0.388 | 0.096 | 1.191 | 0.237 |
| **Hypertension** | 0.204 | 0.491 | 0.033 | 0.415 | 0.679 |
| **Atrial fibrillation** | 0.574 | 0.412 | 0.112 | 1.394 | 0.167 |
| **Dyslipidemia** | -0.064 | 0.404 | -0.013 | -0.158 | 0.408 |

**Supplementary Figure 1.**

Flowchart of patients enrollment and final study sample

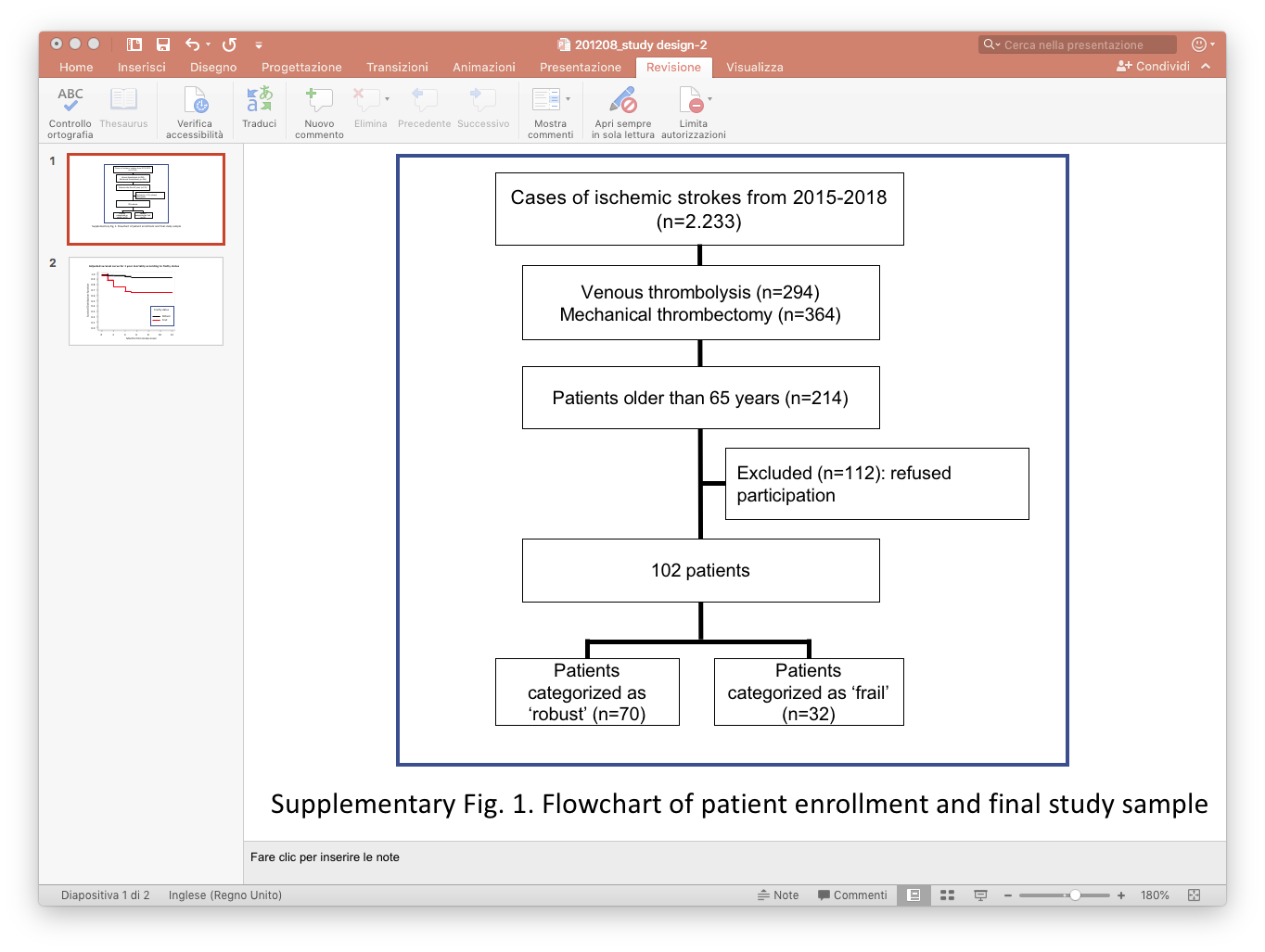

**Calculation of different domains of multidimensional prognostic index.**

The Multidimensional prognostic index was assessed as previously reported (Pilotto et al. 2008). Premorbid functional status was evaluated by the activities of daily living (ADL) score (Katz et al. 1970) and by the instrumental activities of daily living (IADL) score (Lawton et al. 1969) prior stroke. Comorbidity was examined using the cumulative illness rating scale (CIRS) (Linn et al. 1968), while nutritional status was explored with the mini nutritional assessment (MNA) (Guigoz et al. 1999). Cognitive status prior the onset of stroke was categorized as normal (0), reported cognitive impairment with no impact on ADL (0.5) and dementia (1). The Exton–Smith scale (ESS) was used to evaluate the risk of developing pressure sores (Bliss et al. 1966).

The number of drugs used by patients prior stroke and the Cohabitation status, i.e., living with family (with spouse and/or other relatives and/or a caregiver were also recorded. For each domain, a tripartite hierarchy was used, i.e. 0 = no problems, 0.5 = minor problems, and 1 = major problems. The sum of the calculated scores from the eight domains was divided by eight to obtain a final MPI risk score between 0 = no risk and 1 = higher risk of mortality.
